# A global database of COVID-19 vaccinations

**DOI:** 10.1101/2021.03.22.21254100

**Authors:** Edouard Mathieu, Hannah Ritchie, Esteban Ortiz-Ospina, Max Roser, Joe Hasell, Cameron Appel, Charlie Giattino

## Abstract

An effective rollout of vaccinations against COVID-19 offers the most promising prospect of bringing the pandemic to an end. We present the *Our World in Data* COVID-19 vaccination dataset, a global public dataset that tracks the scale and rate of the vaccine rollout across the world. This dataset is updated regularly, and includes data on the total number of vaccinations administered; first and second doses administered; daily vaccination rates; and population-adjusted coverage for all countries for which data is available (138 countries as of 17 March 2021). It will be maintained as the global vaccination campaign continues to progress. This resource aids policymakers and researchers in understanding the rate of current and potential vaccine rollout; the interactions with non-vaccination policy responses; the potential impact of vaccinations on pandemic outcomes such as transmission, morbidity, and mortality; and global inequalities in vaccine access.

## Main

As of 17 March 2021 there have been 2.7 million confirmed deaths and 121 million confirmed cases of SARS-CoV-2 — the virus that causes COVID-19.^1^ Since the beginning of the pandemic and up until today, virus transmission and mortality has been reduced through a range of measures: precautionary actions from individuals including social distancing, wearing facemasks, hand hygiene, and restricting interpersonal contact to outdoor settings; widespread testing to identify individuals infected with the virus; and non-pharmaceutical policy responses from governments – including school and workplace closures; bans on public gatherings; travel restrictions; and stay-at-home orders.^2,3^ Now, with the successful development, evaluation and production of multiple vaccines, governments are turning towards vaccination as an essential solution to the pandemic.

To understand the scale and rate of the vaccine rollout, we need timely, comparable data across countries. The *Our World in Data* COVID-19 vaccination dataset provides a public aggregated global dataset on administered vaccinations. It covers the full period from 13 December 2020 – the date of the first publication of vaccination data – and is being updated regularly ever since. The COVID-19 vaccination dataset is continuously expanding as more countries begin releasing official data on their new national vaccination campaigns. As of 17 March 2021, the dataset covers 138 countries. Our intention is to maintain this database for the foreseeable future and include additional countries as the vaccination campaign starts in a growing number of countries.

This dataset tracks the total number of COVID-19 vaccinations administered by country; broken down by first and second doses (where national data is made available); and derived daily vaccination rates and population-adjusted figures. The combination of these metrics allows users to understand the scale and rate of vaccine rollouts relative to population; compare rollout rates between countries; and assess differences in prioritization for countries with one- and two-dose schedules. This data is compiled from official sources, including health ministries, government reports and official social media accounts.

Our COVID-19 vaccination dataset is already used widely by journalists, policymakers, researchers and the public. The World Health Organization (WHO) relies on this dataset for its official COVID-19 dashboard.^4^ Our dataset is also used by policymakers to benchmark the performance of national vaccination programs across countries. The WHO have also relied on this dataset to understand the inequities in global vaccine access, using this evidence to support calls for greater financial support for COVAX, a global initiative supported by the WHO that is aimed at equitable access to COVID-19 vaccines

Our dataset has been used by all leading media outlets, including the New York Times, the BBC, the Financial Times and The Economist.^5–8^ Global media plays an important role in informing the public during this global pandemic, and it is essential that leading media outlets have timely, transparent and reliable data to present to their audiences. The demonstration of rapid vaccine rollouts across the world, and its potential impacts on transmission and mortality has the potential to shape public attitudes towards vaccinations, reduce vaccine hesitancy, and ultimately lead to an improved response to the pandemic. An effective vaccination response relies on high uptake rates.^9^ As such, our dataset plays a key role in building the public trust that is essential to an effective global response to the COVID-19 pandemic.

Our dataset has been widely-used by the scientific community across multiple disciplines. It has been used to highlight global inequalities in vaccine access, with strong calls to action for the acceleration of financial and policy response efforts to close the existing gaps.^10^ It has been used by researchers to identify countries with particularly effective vaccine rollouts, thereby enabling an analysis of how this was achieved.^11^ These analyses emphasize a range of drivers that explain the large differences across countries that we document: differences in the funding of the development and production of the vaccines; differences in the scheduling and management of vaccinations; differences in public trust and uptake rates; and differentiated responsibilities between national, regional and local level actors.^12^ Health policy researchers have used this dataset to assess differences in vaccine prioritization strategies — for example, which groups should be offered the vaccine first.^13^ Other research groups have combined it with vaccine development data to provide a complete overview of the global vaccine landscape.^14^ It has been used by researchers looking at the role of vaccine hesitancy, and how to design public messaging based on these concerns.^15^

As vaccination campaigns continue to scale, evidence for their effectiveness in reducing transmission, severe disease and death will become increasingly important. Our dataset provides an essential input for epidemiologists who study these questions. Integrating it with other epidemiological data can help researchers evaluate these outcomes.^16,17^ Evidence of positive impacts of vaccination on transmission and mortality can also in turn help to strengthen public trust.

The rest of this article is organized as follows: The Results section presents the key metrics used to track vaccine rollout in this dataset, and the headline results so far. The Discussion section outlines the significance and limitations of this work. The Methods section provides detailed descriptions of the underlying methods and sources used to build this global dataset.

## Results

### Global coverage of COVID-19 vaccination campaigns

The first published reports of COVID-19 vaccinations (outside clinical trials) occurred on 13 December 2020 in the United Kingdom. Our live dataset presents the time-series of vaccinations across the world since then.

To date, 138 countries reported vaccinations, and are included in this dataset. As of 17 March 2021, there have been 400 million doses administered globally. 3% of the world population have received at least one dose of an approved vaccine. This highlights important inequalities in global vaccine access (Figure 1). At the time of writing most high- and middle-income countries have begun vaccination rollouts, but many low-income countries have not (Figure 2). Only 11 countries in Sub-Saharan Africa had started community vaccination.

**Figure 1.**
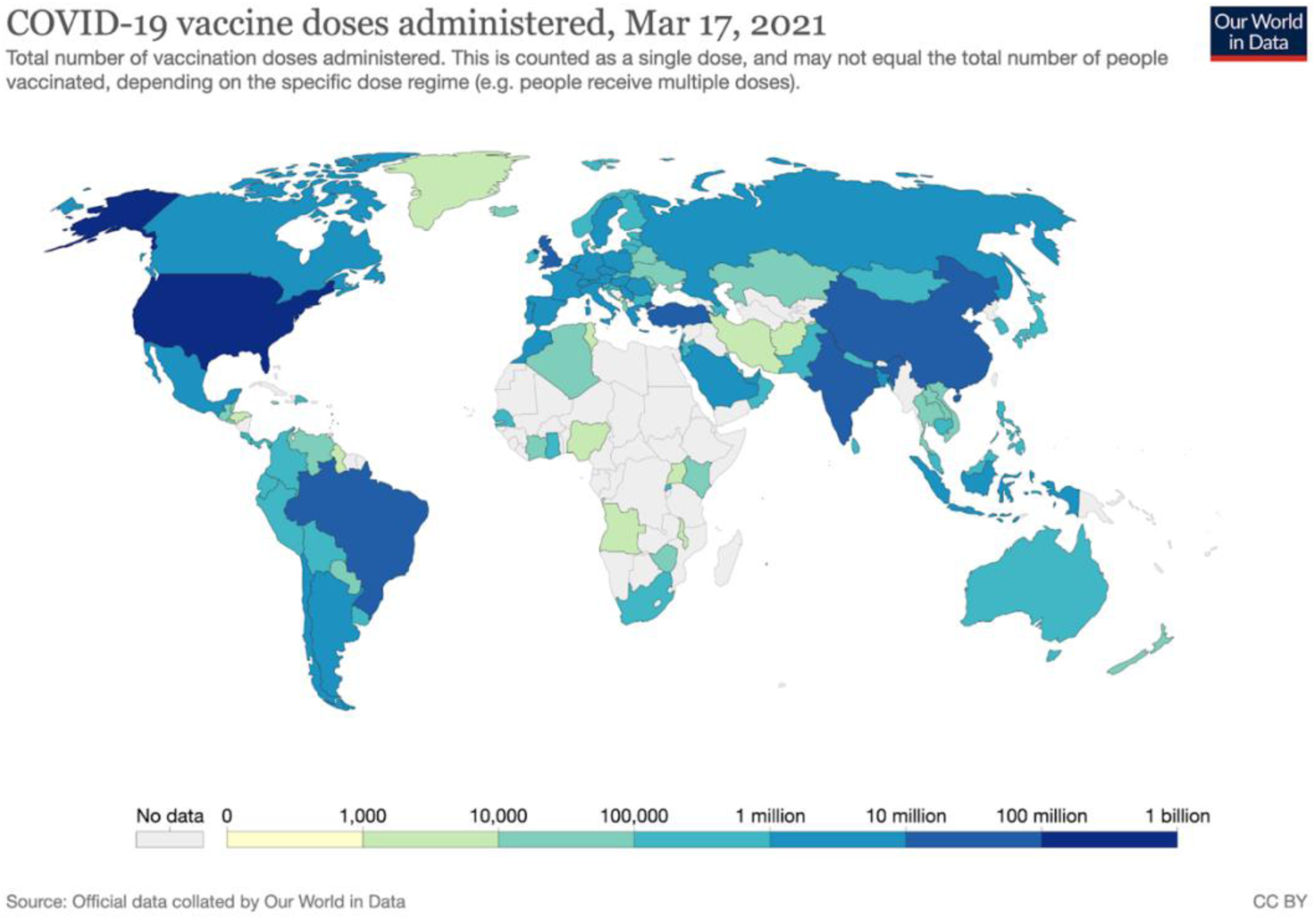
Cumulative number of COVID-19 doses administered by country. This represents the current coverage of the global vaccination rollout.

**Figure 2.**
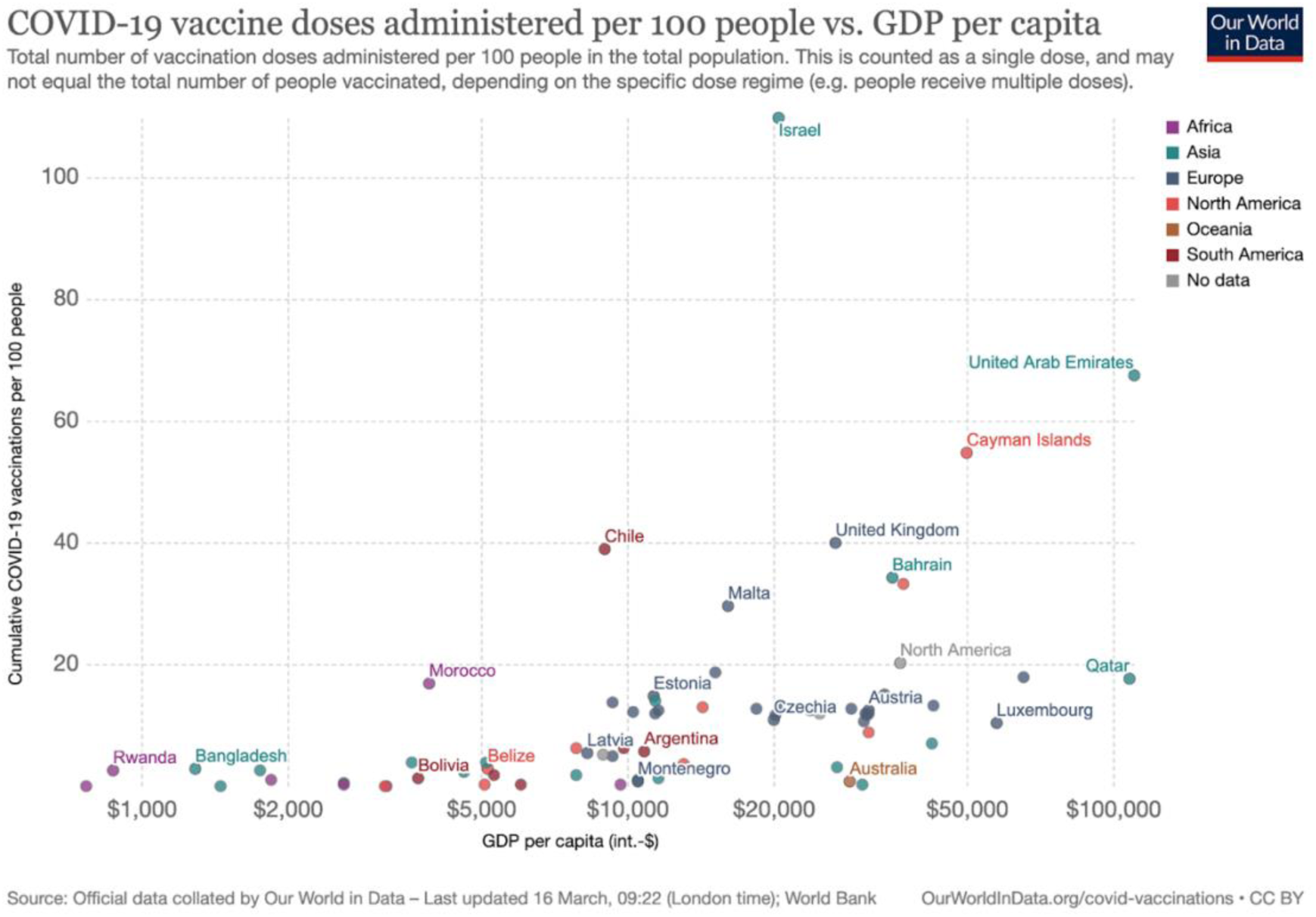
Cumulative number of COVID-19 doses administered per 100 people, measured against gross domestic product (GDP) per capita. GDP per capita is PPP-adjusted, and measured in international-dollars.

### Large differences in vaccination rates between countries

The data reveals large differences in the scale of the vaccine rollout across countries. As of 17 March 2021, we see the cumulative number of doses administered per 100 people range from 110 per 100 in the case of Israel, to 0.01 doses per 100 in countries that have just begun their vaccination campaigns, such as Nigeria, Iran and Vietnam (**Figure 3**).^1^

**Figure 3.**
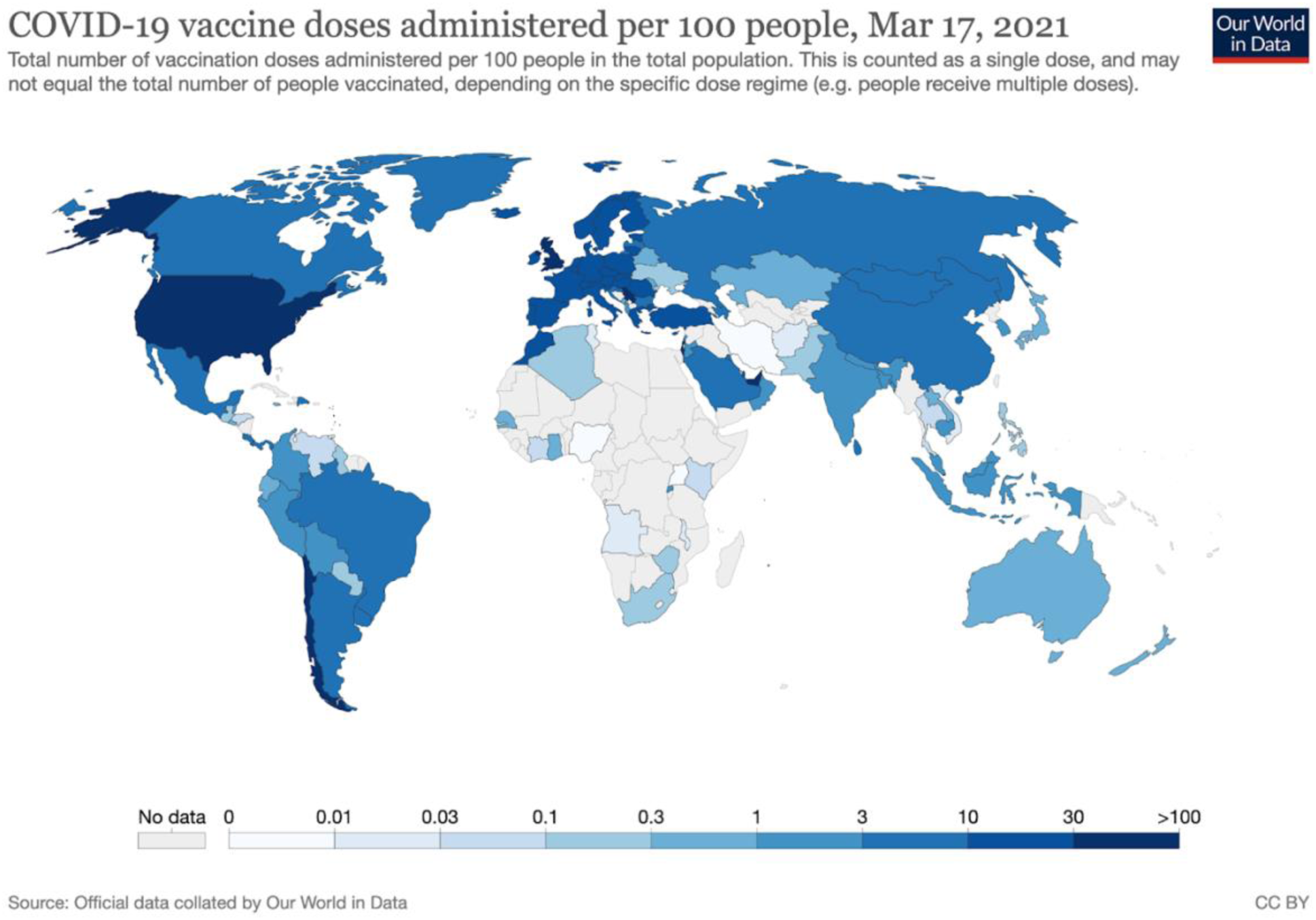
Cumulative vaccine doses administered per 100 people in the total population. Shown is the global situation as of 17 March 2021.

These differences in vaccination coverage are dependent on several factors. First, the date when countries started national vaccination campaigns: the United Kingdom, for example, began community vaccination on 8 December 2020 while other countries have not yet begun their vaccine rollouts.

Second, the rate of vaccinations over time. We see large differences in these rates between countries (**Figure 4a, b**). Israel has received significant attention for the rate of its campaign.^11^ As well as being one of the first countries to begin vaccinations, it also maintained a consistently high rate of vaccinations over time. This is reflected in its steep linear time-series trend of cumulative doses **(Figure 4a)** and consistently high rate of daily doses **(Figure 4b)**. Since the end of December 2020 Israel has averaged a rate of approximately one dose per 100 people per day — more than twice the rate of most countries. Rosen et al. (2021) looked at the contributors to Israel’s success and identified factors such as the organisational and logistical capacity of its community-based healthcare providers; a clear prioritisation framework; and effective outreach efforts to the public as important.^11^ Lee al. (2021) also looked at the factors in Israel’s success, and emphasized the high level of public trust in particular.^12^

**Figure 4.**
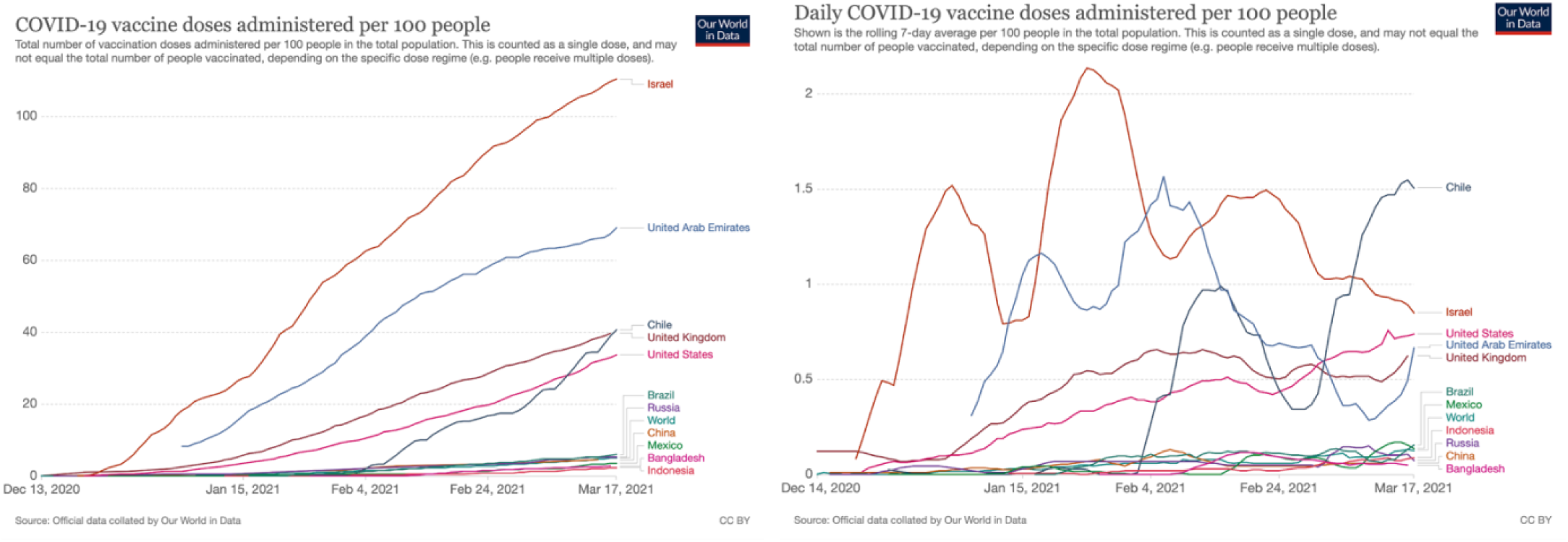
Vaccine doses administered per 100 people: **(a)** shown as the cumulative total per 100 people; and **(b)** daily doses administered per 100 people (7-day smoothed) for select countries.

Most countries which have achieved the fastest vaccine rollouts to date — Israel, United Arab Emirates, United Kingdom, United States, Bahrain, and Chile — are high-income countries. We see from Figure 2 that income matters. But the significant variation at different income levels shows that it’s not the only factor. This can be the basis for research into effective strategies at different income levels.

### Different approaches and prioritization strategies between countries

The *Our World in Data* COVID-19 vaccination dataset allows for comparison of cumulative doses administered, and for a subset of countries, disaggregated data on the number of first and second doses is available. This allows for a comparison of prioritization strategies – a central policy and research question.

Our data highlights large differences in approaches taken by different countries. Some countries — the United Kingdom being the most prominent example — have taken a ‘first dose first’ approach, delaying the delivery of a second vaccine dose to achieve wider single-dose coverage within the total population. This is reflected in the data which shows the share of the total people that have received at least one dose of a COVID-19 vaccine **(Figure 5a)** and the share that have been fully vaccinated **(Figure 5b)**. As of 17 March 2021, 37.2% of the total population had received at least one dose, but only 2.6% had received both doses. Other countries have put greater emphasis on giving two doses to a smaller share of the population. In these countries a large share of those who received the first dose have already received the second dose: In Israel, 59.5% had received at least one dose, and 50.9% had been fully vaccinated. In the United States, 22% had received one dose and 12% had been fully vaccinated.

**Figure 5.**
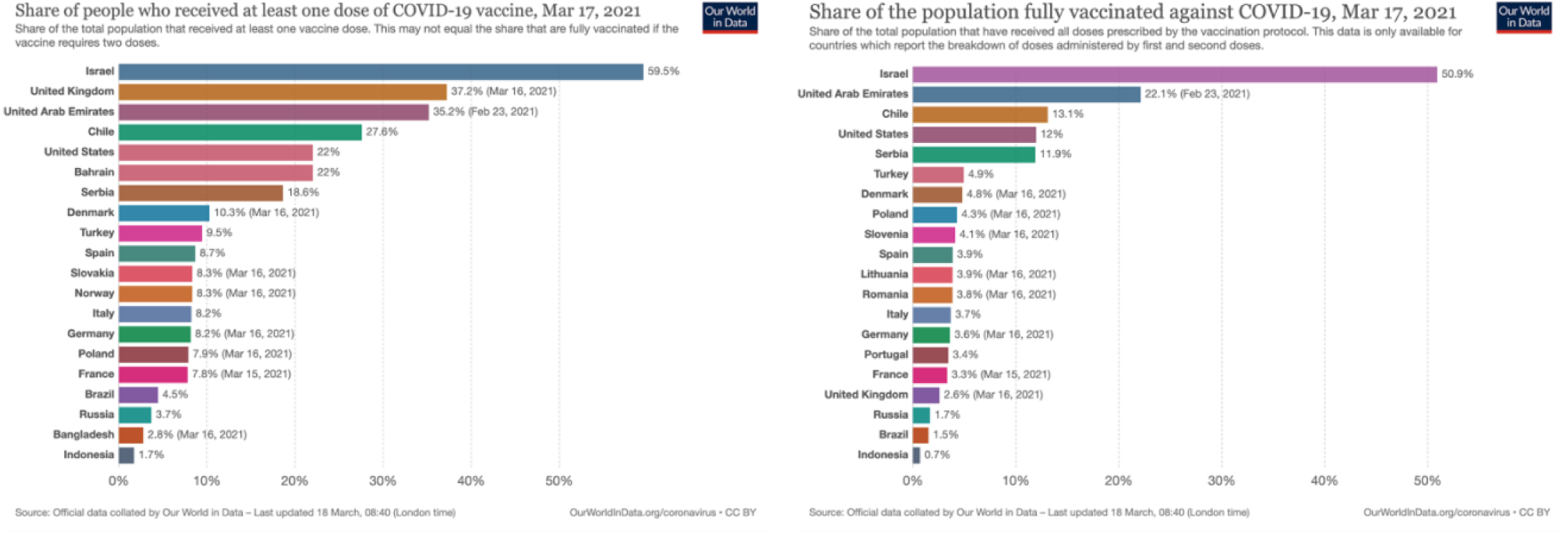
Share of the total population that have: **(a)** received at least one dose of the COVID-19 vaccine; and **(b)** been fully vaccinated against COVID-19, for select countries.

Differences in prioritization have received particular attention in Europe. The ‘first dose first’ approach favoured by the United Kingdom has been frequently contrasted with the approach of many countries in the European Union.^18–20^ This is reflected in our dataset: Germany, for example, has administered a first dose to a much smaller share of the population (8.1%) but has fully vaccinated a higher share than the UK (3.6%). While the share of the population that received a first dose in Germany was much lower than in the UK, the share who received a second dose is actually higher.

## Discussion

The rapid development, testing and manufacturing of multiple effective vaccines against SARS-CoV-2 was a ground-breaking achievement in 2020. Never before in history has a vaccination campaign started so very soon after a new pathogen was identified. In many cases it took many years or decades until a vaccine was developed (Figure 6). In the case of COVID-19 scientists have developed several, highly efficacious vaccines within the same year. The question now is whether the global rollout of the vaccine can match the speed with which the vaccine was developed: whether they can be administered quickly and equitably across the world.

**Figure 6.**
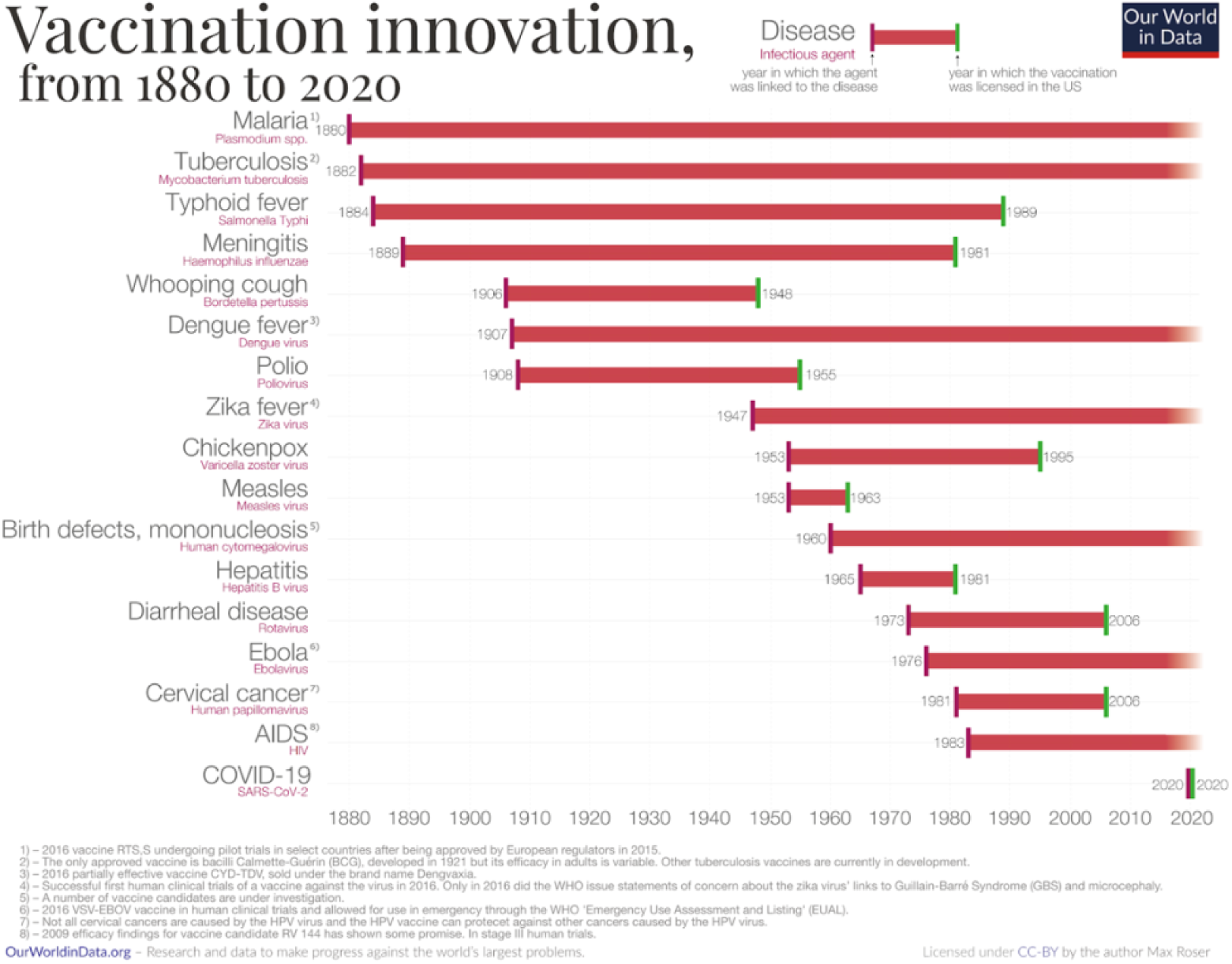
Timeline of innovation in the development of vaccines. Each bar begins in the year in which the pathogenic agent was first linked to the disease and the bar ends in the year in which a vaccination against that pathogen was licensed in the US.

To do this, governments and public health officials need to understand the most effective approaches to mass vaccination rollouts and prioritize the administration in a way that minimizes morbidity and mortality from the disease. The dataset that we present here provides an essential resource to support this. It allows analysts to track vaccinations over time in a specific country, and also to compare vaccination rates and prioritization strategies across countries. Combined with epidemiological data, it is a vital input for researchers to understand how vaccination affects the transmission and health outcomes of COVID-19.

Communicating such research on the efficacy of vaccines is in turn essential for building public trust and reducing vaccine hesitancy.^21,22^ Kreps et al. (2021) found that perceived vaccine efficacy was the strongest predictor of COVID-19 vaccine uptake in the United States.^23^ Sherman et al. (2021) studied people’s willingness to be vaccinated against COVID-19 and their attitude towards vaccines.^24^ The researchers documented that the effectiveness of vaccines in reducing disease is a particularly important argument.

It is important to highlight what we do not try to achieve with this dataset collection. We only collect data on doses administered — we do not include data on the number of doses manufactured, ordered or delivered. In the collection of data on administered doses we also do not audit official reports beyond technical validation (see Methods section). And we also do not attempt to assess vaccine effectiveness or the impacts of vaccinations on pandemic outcomes. This is beyond the scope of this resource.

If we want to understand anything about vaccines – effectiveness, policy responses, perceptions – then we need to know how many vaccines have been administered. Our dataset fills this gap.

## Methods

In this section we first provide a description of the metrics made available in this dataset, followed by information on how this data was collected. Finally, we describe how it is published as a complete, open-access dataset and how it can be explored via our online web application.

### Metrics included in this dataset

The metrics included in this dataset are a combination of original metrics reported by official sources, and derived metrics calculated by *Our World in Data*.

The nine metrics included in this dataset are the following.

1. **Total doses administered**. This is a count of all doses given. Since several vaccines require multiple doses this count may be higher than the total number of people vaccinated.
2. **Total doses administered per 100 people**. This is ‘Total doses administered’ per 100 people of the total population.
3. **Daily vaccinations**. If official sources provide daily updates of vaccinations administered, this is included.
4. **Rolling average of daily vaccinations over 7 days**. For countries that don’t report data on a daily basis, we assume that the number of administered doses was the same on all days over any periods in which no daily data was reported. This produces a complete series of daily figures, which is then averaged over a rolling 7-day window.
5. **Daily doses administered per million people**. This is the rolling average of daily vaccinations over 7 days, per million people within the total population.
6. **Number of people that have received at least one vaccine dose**. Depending on the vaccine schedule (a one or two-dose vaccine), this may or may not match the number of people fully vaccinated. If a person receives the first dose of a 2-dose vaccine, this metric goes up by one. If they receive the second dose, the metric stays the same.
7. **Share of the total population that have received at least one vaccine dose**. This is the number of people that have received at least one vaccine dose per 100 people in the total population.
8. **Number of people that are fully vaccinated**. This is the total number of people who received all doses prescribed by the vaccination protocol. If a person receives the first dose of a two-dose vaccine, this metric stays the same. If they receive the second dose, the metric goes up by one.
9. **Share of the total population that are fully vaccinated**. This is the number of people that have received all doses prescribed by the vaccination protocol per 100 people in the total population.

Not all metrics are available for all countries. The availability is dependent on the granularity of reporting provided by the official sources. For example, not all countries provide a breakdown of doses administered by first or second doses. In this case, only the total number of administered doses can be provided for this country.

### Data collection methods

Raw data on vaccination doses administered is collected through a combination of manual and automated means. This collection process differs by country, but can be broadly defined by three methods.

Firstly, for a number of countries, figures reported in official sources — including government websites, health ministries, dedicated dashboards, and social media accounts of national authorities — are recorded manually as they are released.

Secondly, where official sources release vaccination figures in a consistent, machine-readable format, or where structured data is published at a stable location, we have automated the data collection via Python scripts that we execute every day. These automated scripts are made available in our GitHub repository (https://github.com/owid/covid-19-data/blob/master/scripts/scripts/vaccinations/automations/). These are regularly audited for technical bugs to ensure data validity (see ‘Technical Validation’, below).

Third, in some instances – where national data is not made available in machine-readable format by official sources, but is aggregated by third-party sources – we source data from non-official publishers (e.g. https://covid19tracker.ca/vaccinationtracker.html for Canada). These are also regularly audited for accuracy against the original official sources.

### Calculating derived metrics

Derived metrics are calculated from the raw official counts in two ways.

1. Population-adjusted metrics. This normalizes total doses, first doses, and second doses to their counts per 100 people within the total population. This allows users to compare the pace and scope of the vaccination rollout across countries. Vaccinations administered per 100 people are calculated by dividing administered doses by total population figures, sourced from the latest revision of the United Nations World Population Prospects.^25^ The exact population values used in these calculations are also provided in our GitHub repository (https://github.com/owid/covid-19-data/blob/master/scripts/input/un/population_2020.csv).

2. Daily rolling averages. Not all countries report figures at a daily frequency. In order to facilitate cross-country comparisons over time, we therefore derive a ‘smoothed’ daily vaccination series calculated as the seven-day moving average. It is calculated as the right-aligned rolling seven-day average of a complete series of daily changes. For countries for which no complete series of daily changes is available from our source, we derive it by linearly interpolating the cumulative totals. The exact code used to derive the 7-day moving average is available online (see ‘Code Availability’, below).

### Criteria for inclusion and coverage

To be included in this dataset, countries must provide at least one data point on the number of vaccine doses administered via a trusted country-specific source such as a health ministry, government report or official national account. Reports on the number of vaccine doses ordered or distributed are not included; this dataset only includes doses administered. Vaccinations administered in clinical trials are also not included.

As of 17 March 2021, the vaccination dataset covers 138 countries. This coverage will continue to expand as more countries begin vaccination campaigns.

### Technical validation

The *Our World in Data* COVID-19 vaccination database represents a collation of publicly available data published by official sources. The main quality concern for the database itself is whether it represents an accurate record of the official data. We employ several strategies to ensure that this is the case.

First, all automated collection of data, whether obtained from official channels or non-official publishers, is subject to initial manual verification when it is added to our database for the first time.

Second, we employ a range of data validation processes, both for our manual and automated time series. We continually check for invalid figures such as negative or illogical values, out-of-sequence dates, implausible changes in time-series data, invalid population-adjusted values, and for each country we test for abrupt changes in vaccination rates.

Third, our vaccination data is viewed and used by millions of people every day — either through direct usage or third-party usage (such as news reports, social media and other sharing channels). This includes employees of health ministries, researchers, journalists, and policymakers from across the world. We receive large amounts of feedback from this user base. This provides a final ‘crowd-sourced’ verification method that has been shown to be effective in highlighting any discrepancies to official data sources.

### Data access and publication

Vaccination data is updated daily and is made available via two channels. Firstly, all data and scripts used for data collection are published and updated in our public GitHub repository (https://github.com/owid/covid-19-data/tree/master/public/data/vaccinations). This provides a transparent resource for users to download the data in CSV and JSON formats; replicate the data collection and metric derivation process; and monitor any changes or additions to this process.

To allow journalists, researchers, policymakers and the public to understand the evolution of the global COVID-19 vaccination rollout, we make all of this data explorable at our online web publication (ourworldindata.org/covid-vaccinations). There we provide interactive visualizations of all available vaccination metrics to allow users to track and compare the vaccination campaigns around the world. These interactive visualizations are built with our custom visualization tool – the *Our World in Data Grapher* – and are made available open-access. They are updated daily in sync with updates in our GitHub repository.

## Data Availability

A live version of the vaccination dataset and documentation are available in a public GitHub repository at https://github.com/owid/covid-19-data/tree/master/public/data/vaccinations. This data can be downloaded in CSV and JSON formats.

## Code availability

Our scripts for data collection, processing, and transformation, are available for inspection in the public GitHub repository that hosts our data (https://github.com/owid/covid-19-data/tree/master/scripts/scripts/vaccinations).

## Footnotes

1 As discussed below, the number of doses can exceed the number of people due to multiple dose vaccination programs.

